# The Peak of COVID-19 in India

**DOI:** 10.1101/2020.09.17.20197087

**Authors:** Suryakant Yadav, Pawan Kumar Yadav

## Abstract

**Introduction:** Following the USA, India ranks the second position in the world for COVID-19 cases with the highest number of daily confirmed cases since September 2020. The peak of COVID-19 cases is the most warranted feature for understanding the curvature of COVID-19 cases.

**Aim:** This study aims to analyse the growth rates of the daily confirmed cases and to provide an expected count of the peak of daily confirmed cases.

**Data:** We retrieved data from an Application Programming Interface portal https://www.covid19india.org/ which is open access and publicly available.

**Methods:** Exponential model was applied to estimate the growth rates of daily confirmed cases. The estimated growth rates were used for calculating the doubling time. The Lotka-Euler method was applied to calculate the effective reproduction rate. SARIMA model was developed for the growth rates to predict daily confirmed cases.

**Results:** Results show the best fit of the exponential model over the daily confirmed cases. The growth rates estimated from the exponential model shows an unsteady, modest decline. Doubling time shows a linear increase. The effective reproduction rate declined from 3.6 persons in the third week of March 2020 to 1.14 persons at the end of August 2020 and 1.12 persons in the mid-September 2020. The diagnosis of the developed SARIMA model confirmed no trends in the residuals, no outliers, and nearly constant variance. The forecast suggests the peak value of daily confirmed cases would waver around 105,000 counts in the third week of September 2020. The cumulative COVID-19 cases would account for approximately 105 lakhs at the end of December 2020.

**Conclusion:** The exponential model unravels a shift and a modest decline in the growth of daily confirmed cases. The trends in *R*(*t*) show analogue with the trends in growth rates of daily confirmed cases. The study shows that the SARIMA model is suitable for projecting daily confirmed cases. The results shed light on the understanding of the trends and epidemiological stage of COVID-19 disease, in the cognisance of the peak.

**Contribution:** This study based on moments of the distribution of the daily confirmed cases of COVID-19 disease unravels the uncertainty about the peak and curvature of COVID-19 disease.

**Research Highlights:**

1. The exponential model is the best fit over daily confirmed COVID-19 cases.
2. The trends in growth rates of daily confirmed cases show a swift decline during the five-and-a-half months period since April 2020.
3. The effective reproduction rate in India declined from 3.6 persons in the third week of March 2020 to 1.14 persons at the end of August 2020 and 1.12 persons in the mid of September 2020.
4. The forecast reveals that the peak of daily confirmed cases wavers at approximately 105,000 cases since the third week of September 2020.
5. The *R*(*t*) value would be equal to one in the first week of December 2020.
6. The cumulative confirmed cases of COVID-19 in India would account to approximately 105 lakhs at the end of December 2020.

## Introduction

In late December 2019, for the first-time pneumonia cases with an unknown cause were reported in Wuhan (China). In January 2020, the cause of those pneumonia cases was identified as a new type of coronavirus (Zhu et al., 2020). On 12 January 2020, the official name of the coronavirus as COVID-19 (stand for Coronavirus disease 2019) for the disease as well as SARS-CoV-2 (severe acute respiratory syndrome coronavirus 2) for the virus was given by World Health Organisation (WHO, 2020).

Coronavirus disease of 2019 (COVID-19) has shown an unprecedented increase worldwide. Following the United States of America (USA), India ranks the second position in the world with cumulate COVID-19 cases at slightly more than 51 lakhs on 24:00 PM 16 September 2020. India has recently surpassed Brazil in the number of COVID-19 cases in the first week of September 2020. It is foreseeable that India can easily surpass the USA as still the peak of COVID-19 cases in India is yet not seen. Also, the growth rates of COVID-19 cases do not show a dramatic and unexceptional decline in near time.

While the USA has experienced the second wave of COVID-19 cases, Brazil has witnessed the peak of COVID-19 cases since the end of July 2020 and after that, has shown a swift decrease in the number of COVID-19 cases. Other countries such as Russia, Peru, and South Africa have exhibited the peak of COVID-19 cases with the cumulative number of COVID-19 cases smaller than what the top three countries have exhibited during the same time (JHU CSSE 2020). Following the unprecedented ‘stay-at-home’ national policies, Europe and many other countries show receding COVID-19 pandemic recently (Vokó and Pitter, 2020). In view of these trends, India shows a different predicament. Unlike other countries, India is the only country showing upsurge of COVID-19 cases without showing a peak.

The peak value of daily confirmed COVID-19 cases in the USA and Brazil was recorded at 77,255 and 69,074, respectively. However, India surpassed the USA in the daily confirmed COVID-19 cases on 1 September 2020 and breached the mark of 95,000 on 16 September 2020. In seven months since the first case on 31 January 2020, the daily confirmed COVID-19 cases have reached to the highest count in the world. The rapid rise of daily confirmed cases in India indicates an exponential growth of COVID-19 cases. This upsurge in COVID-19 cases is an alarming situation as most of the governing body guidelines have been in the best hope to attain the most awaited peak as early as possible.

India has taken note of the rise in COVID-19 cases. In view of that, the Government of India (GOI) has implemented lockdown from 25 March 2020 in many phases. For different phases of lockdowns, GOI has issued many guidelines from time to time intending to maintain social distancing. From June 2020, the GOI has also implemented unlocks with proper guidelines. In the absence of vaccine, medicine and drugs, these non-pharmaceutical interventions are the precautionary measures in a bid to flatten the curve of COVID-19 cases (Atangana, 2020) accompanied by low growth rates as well as low effective reproduction rates. The success of non-pharmaceutical interventions is evident in many countries while encountering the spread of SARS-CoV-2 virus. The importance of non-pharmaceutical interventions, including isolation and wearing masks, to control the disease transmissibility is considered as successful among the Italian population despite a toll of deaths (Chintalapudi et al., 2020).

Effective reproduction rate (*R*(*t*)) is an essential epidemiological measure of an epidemic or pandemic. For COVID-19 disease, *R*(*t*) measures the spread of SARS-CoV-2 from a primary infectious person to secondary infectious persons at a time ‘*t*’. *R*(*t*) below a value of one is warranted to contain the spread of disease. Having achieved that, the epidemics shrink (Kupferschmidt, 2020). While vaccines are still under the process of development, social distancing, isolation, extensive testing, and quarantining of confirmed infected cases remain the most effective measures to contain the pandemic (Tsay et al., 2020; Liu et al., 2020).

The rise of confirmed cases has been evident in India. Therefore, authorities and academicians are looking forward to the peak of the confirmed cases as the best outcome of non-pharmaceutical interventions in the absence of vaccine, drugs, and medicine.

The objectives of the study are to:

1. analyse the growth rates of the confirmed cases of COVID-19 in India,
2. provide an expected count of the peak of confirmed cases and a possible track of confirmed cases.

## Data and Methods

### Data

We retrieved data from an Application Programming Interface (API) portal https://www.covid19india.org/ which is open access and publicly available (COVID19-India API 2020). Data on COVID-19 cases on this portal is updated from state bulletins, official handles, PBI, Press Trust of India (PTI), and Asian News International (ANI) reports. This study uses data from the API portal ‘covid19india.org’ because of the two advantages. The first is that it provides time-series data in a portable ‘*.csv’ files, and the second is that it provides data up to the district level of India in the same format.

This ‘covid19india.org’ portal provides data on confirmed, active, recovered, and death cases, on a daily basis, from 30/01/2020. We retrieved data between the dates 30/01/2020 and 16/09/2020, for analysing data for seven-and-a-half months period. The first case in India was reported on 30/01/2020. We performed analyses for the periods rolling from 15/03/2020 to 31/08/2020, a period of five-and-a-half months. The daily confirmed cases for the period rolling from 01/09/2020 to 16/09/2020 is used for validating for the next 16 days.

This period of five-and-a-half months are consisting of the time-intervals as lockdowns and unlocks. These are Lockdown 1.0: 25/03/2020–14/04/2020, Lockdown 2.0: 15/04/2020–03/05/2020, Lockdown 3.0: 04/05/2020–17/05/2020, Lockdown 4.0: 18/05/2020–31/05/2020, Unlock 1.0: 1/06/2020–30/06/2020, Unlock 2.0: 1/07/2020– 31/07/2020 and Unlock 3.0: 1/08/2020–31/08/2020. The Unlock 4.0 starts from 01/09/2020.

The results in figures are shown with these time-intervals. The daily confirmed cases, adjusted R-square, *R*(*t*), and growth rates are updated up to the latest possible date.

## Methods

### Exponential model and doubling time

The plot of confirmed cases of COVID-19 disease over time suggests an exponential model. The exponential model was applied with an intercept and also without an intercept. However, parameter estimates for the intercept converged to zero, and other shape and scale parameters converged to a significant value. One advantage of the exponential model is that it does provides time-invariant parameter estimates for the time-series data available for India. The haphazard trends in parameter estimates by states of India do not allow projections. Data at a lower geographical level in India is still a limitation for robust analysis.

The exponential model can take the form of growth and decay model depending upon growth rate greater than zero and smaller than zero, respectively. Exponential growth (decay) model is preferred for the following two reasons. First of all, the data has not shown any peak given the trends into consideration, and second, the acceleration or deceleration of the growth of confirmed cases is not explicit.

The exponential model is expressed as

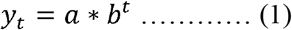

where *t* is time or date, *y_t_* is the number of daily confirmed cases at time *t, a* is constant, *b* is the acceleration or deceleration of the fit of the daily confirmed cases, and *b = e^r^* with r as the growth rate of daily confirmed cases.

Non-linear estimation method was applied on daily confirmed cases between 31 January 2020 and rolling from 15 March 2020 to 31 August 2020 to estimate of the value of model parameters *‘b’* and *‘a’* to get the best fit until each date. The estimated values of parameters ‘b’ and ‘a’ are time-invariant. The natural logarithm of the estimated value of parameter *b* from the exponential model gives the estimate of growth rate ‘*r*’ of daily confirmed cases.

The doubling time (*T_d_*) (Galvani et al. 2003) of COVID-19 cases using the growth rate of daily confirmed cases is expressed as

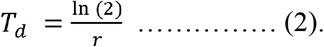

### Effective reproduction rate

The Lotka-Euler equation (Wallinga and Lipsitch 2007) was applied to estimate the effective reproduction rate based on the growth rate of daily confirmed cases of COVID-19. The Lotka-Euler equation is expressed as

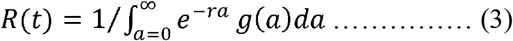

where, *R*(*t*) is the basic reproduction rate, denoted by *R_O_*, in the initial period ∼5 to ∼14 days of the spread of infectious disease, and is the effective reproduction rate, denoted by *R*(*t*), at a time *‘t’* of the outbreak of the disease; *r* is the growth rate of daily confirmed cases; g(a) is the probability density function of generation (serial) interval which is the time-span from a primary infectious person to generate secondary infectious persons in the time interval [*a, a + da*] (Ma 2020).

Mathematically, *R*(*t*) depends on two statistics: the growth rate of daily cases and serial interval distribution. In the lack of duration of serial intervals in the observed duration of the infectious period, hence, serial interval distribution, serial interval distribution applicable for China between 21 January 2020 and 8 February 2020 (Du et al. 2020) was applied for India. Du et al. (2020) tested models such as Normal, Lognormal, Gamma and Weibull distribution for 469 reported transmission events. They provided the shape and scale of these serial interval distributions. Of these models, the Gamma distribution shows the best fit of serial interval data in China. The Gamma distribution with the shape and scale equal to 1.46 and 0.78, respectively, (Du et al. 2020: 4) is useful for India.

### Projection of confirmed cases

Auto Regressive Integrated Moving Average (ARIMA) was applied to the growth rate of daily confirmed cases of COVID-19 (Singh et al. 2020, Ceylan 2020) to predict daily cases until 28 February 2021. ARIMA (p, d, q), where p is the order of autoregression, d is the degree of difference, q is the order of moving average, is expressed as

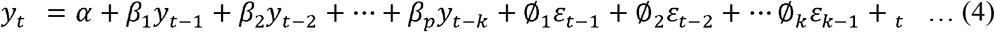

where *y_t_* is the differenced series of confirmed cases, *y_t-k_* is the lagged values of *y_t_* of order *k, _t_* is white noise, *ε_k-1_* is the lagged errors of order *k, ∝* is an intercept term, *β* is an autoregressive parameter, and ∅ is a moving average parameter.

An ARIMA model is efficient in dealing at non-stationary and stationary time series. The SARIMA for time series is referred to as an ARIMA (p, d, q) (P, D, Q)[m] where (P, D, Q) represents the (p, d, q) for the seasonal part of the time series, and m refers to the number of observations per cycle. The accuracy of the SARIMA model is measured by root mean square error (RMSE) and mean absolute percentage error (MAPE).

## Assumptions

The study assumes:

1. the trends in the future would be analogous to that of the trends in the past
2. the seasonality in the trends in the past remains the same in the future,
3. the number of testing and the test positivity ratio does not change dramatically.

## Results

### Exponential model

Figure 1 shows the non-linear fits based on the Equation (1), which is an exponential function, over the daily confirmed cases until two dates at the end of May and August 2020. The daily confirmed COVID-19 cases are shown until 16 September 2020. These non-linear fits (deep-sky-blue (dark and light) coloured long-dash line) vividly shows the exponential increase in daily confirmed cases. An interesting finding to ponder is the shift of the non-linear fit over time. The shift in the exponential fit indicates a slowdown in the increase of daily confirmed cases. Although the number of daily confirmed cases has been increasing, the cases seem lesser than expected counts of cases based on the exponential model.

**Figure 1:**
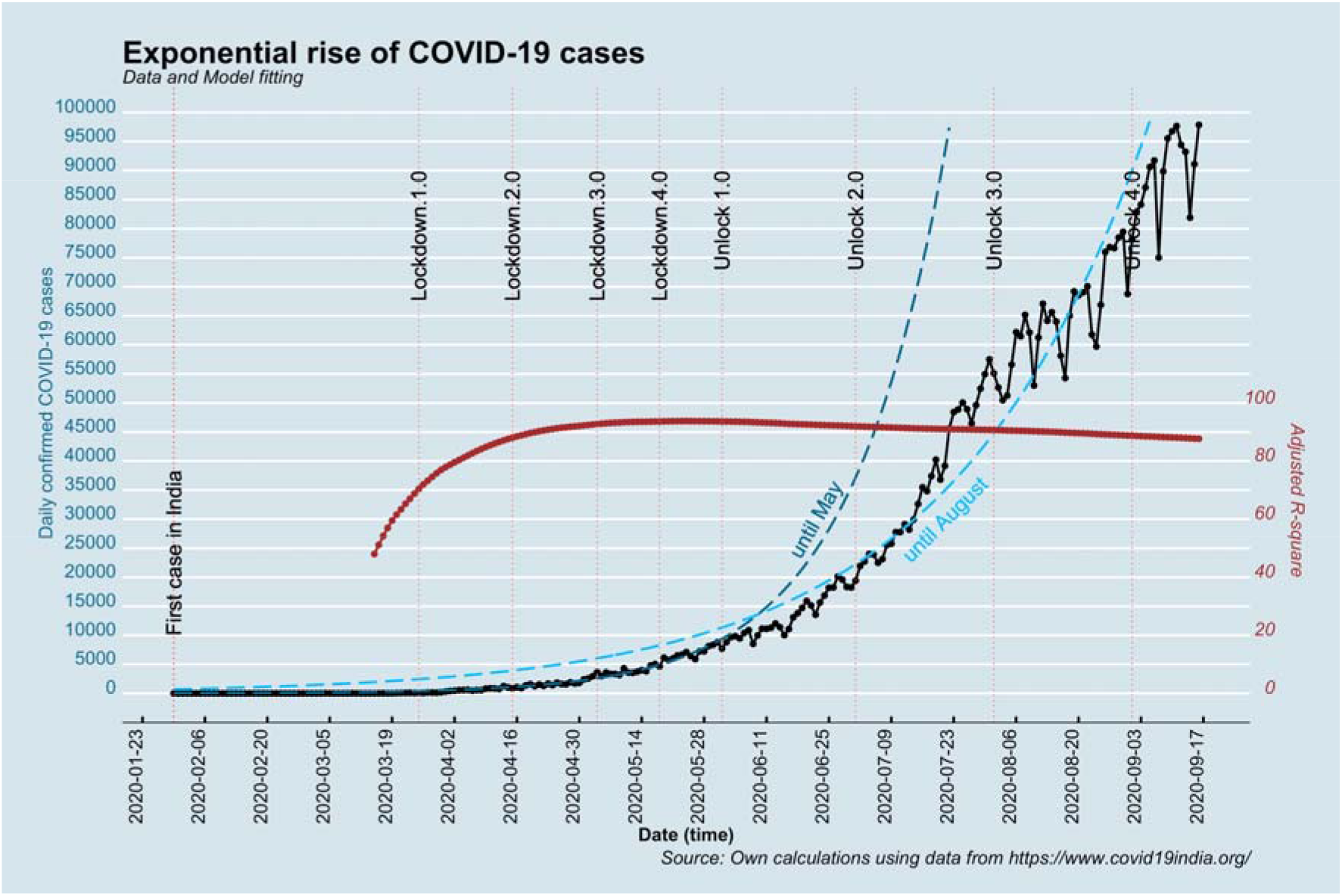
Plot of exponential fits over daily confirmed cases, India.

Nonetheless, the burgeoning daily confirmed cases are supported by the fact that the adjusted R-square values (brown coloured connected circles) of the exponential fit is more than 85 per cent from May to August 2020. It is noteworthy that the fit of the exponential model over the data has become stronger since May when compared from mid-March to April.

### Growth rates and doubling time

Figure 2 displays growth rates and doubling time calculated from the estimate of parameter ‘b’ of the exponential model during the period of five-and-a-half months period from mid-March to August 2020. It is shown until 16 September 2020. The parameter ‘b’ or the slope of the exponential model explains the acceleration or deceleration of daily confirmed cases. The shift in the fit of the exponential model is an indication of a change in the slope, i.e., a change in the growth rates (*r*) of daily confirmed cases.

**Figure 2:**
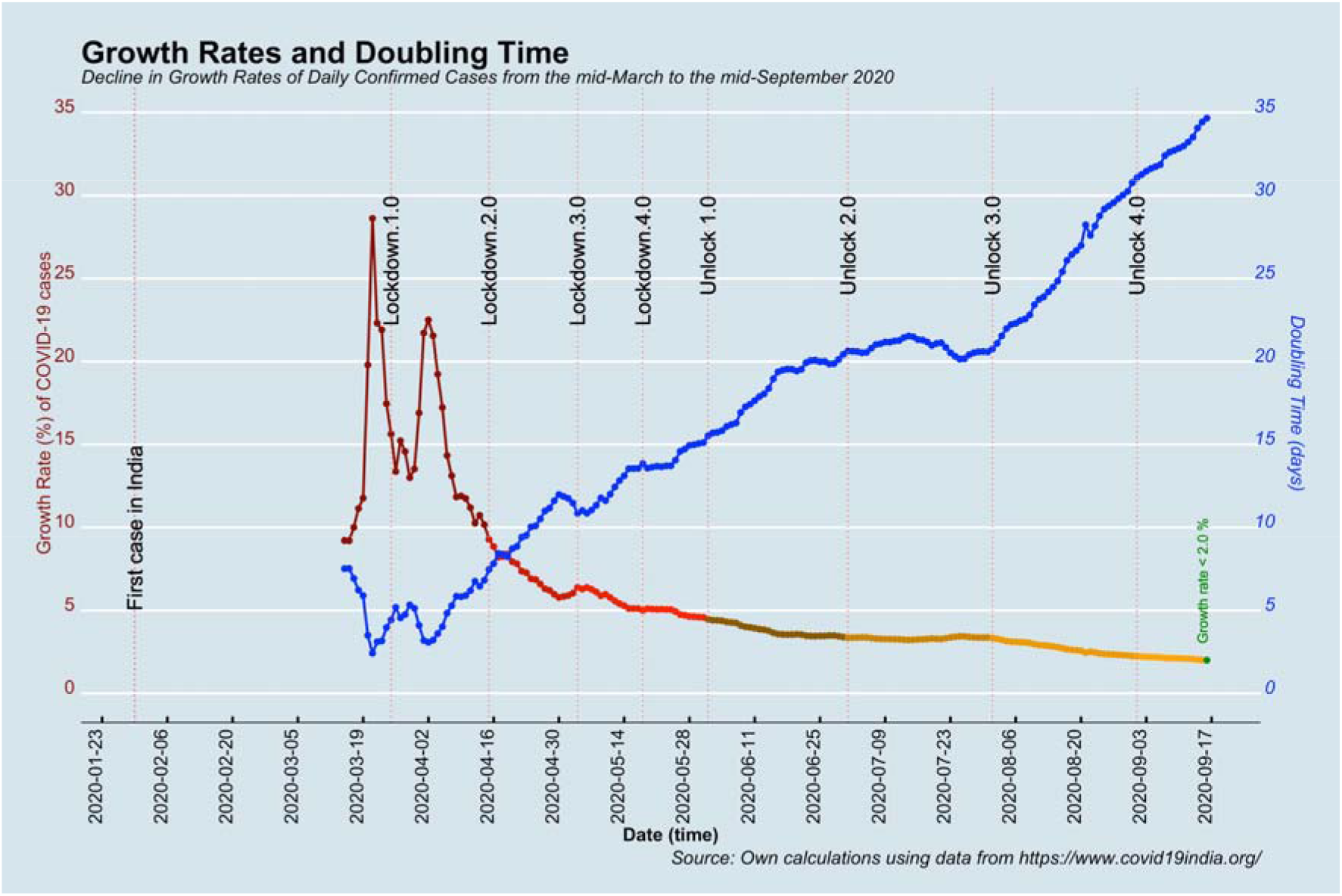
Trends in growth rates of daily confirmed cases and doubling time of cumulative COVID-19 cases.

The trends in the growth rates of daily confirmed cases show high and non-steady values of growth rates with spikes in the latter half of March and April. The highest value of growth rate was 28 per cent in the third week of March 2020. The growth rates declined rapidly since the first week of April, and down the line, it declined to approximately a value of 5.5 per cent in the last week of April. However, at this end, the growth rates showed an abrupt increase. In the first week of May 2020, it increased to 6.4 per cent. This rise in growth rates was susceptible to return of migrants from metro cities to their homeland mostly in rural areas. The rise in growth rates has affected the trends in growth rates as its gradients ramped up. Also, a stagnancy in growth rates is evident, and it declined slowly to 5 per cent in the last week of May 2020. The growth rates declined, however, slowly in April and May.

Altogether, the lockdowns were successful in lowering the growth rates; however, the pace of decline was slow. The 5 per cent growth rate at the end of May were high enough to push the daily confirmed cases to a new height.

The trends in growth rates of daily confirmed cases show a rapid decline in June 2020. By the end of June, the growth rate values declined rapidly to approximately 3 per cent, and the doubling time increased from 10 to 15 days. Nonetheless, 3 per cent growth rate is a high growth rate as the daily confirmed cases confirm an exponential increase. However, the growth rates did not decline in next month, and it was almost stagnant in the subsequent month of July 2020. Instead, there was a slight increase in the growth rates, and that added the stacks of daily confirmed cases in July. At that nearly-constant growth rate, the number of daily confirmed cases rose from approximately 20,000 to 60,000 in July. At a doubling time of approximately 20 days, the cumulative confirmed cases were approximately 17 lakhs at the end of July. Ten lakhs more confirmed COVID-19 cases were added during July in addition to approximately four lakhs confirmed cases during June. In the backdrop, there is a strong base for the exponential growth rate to manifold increase in daily confirmed cases. With such a big base, a small positive exponential growth rate is sufficient to pile up daily confirmed cases. It would keep the peak of confirmed cases faraway. By the end of July 2020, India’s position regarding COVID-19 pandemic was entirely different what could have been just one month ago.

The growth rates rollbacked on the path of smooth decline in August 2020. The growth rates declined swiftly from 3.3 per cent to 2.2 per cent. Despite this decline in growth rates, the daily confirmed cases kept on burgeoning in this month because of a large base. The doubling time increased from 20 days to 31 days. Accordingly, the cumulative COVID-19 cases increased by twofold and added approximately 19.4 lakhs in August. India shows the doubling rate of 31 days after seven months at a stockpile of daily confirmed cases. By the end of August, the count of daily confirmed cases increased to 68,767.

India has taken seven months to reach a low growth rate. The delay in reaching a low growth rate has a cost in terms of a high peak and a large cumulative confirmed case. India can circumvent this delay by having a low growth rate of value one per cent sooner rather than later; however, this delay is inevitable. At the growth rate of one per cent, the exponential model becomes a function of time (days) and the parameter estimate ‘*a*’, and then, the daily confirmed cases are not growing exponentially. Nonetheless, daily confirmed cases would be multiplicative of time, but it cannot have a snowballing effect. The peak of the daily confirmed cases would appear for a growth rate between one and two per cent. Since the middle of September, India is passing through a crucial time when there is a possibility of observing the peak of daily confirmed cases.

### Tracking effective reproductive rates

The effective reproduction rate *R*(*t*) is one of the epidemiological measures to understand the spread of COVID-19 disease. Figure 3 shows the trends in *R*(*t*) from the latter half of March to August 2020. It is shown until 16 September 2020.

**Figure 3:**
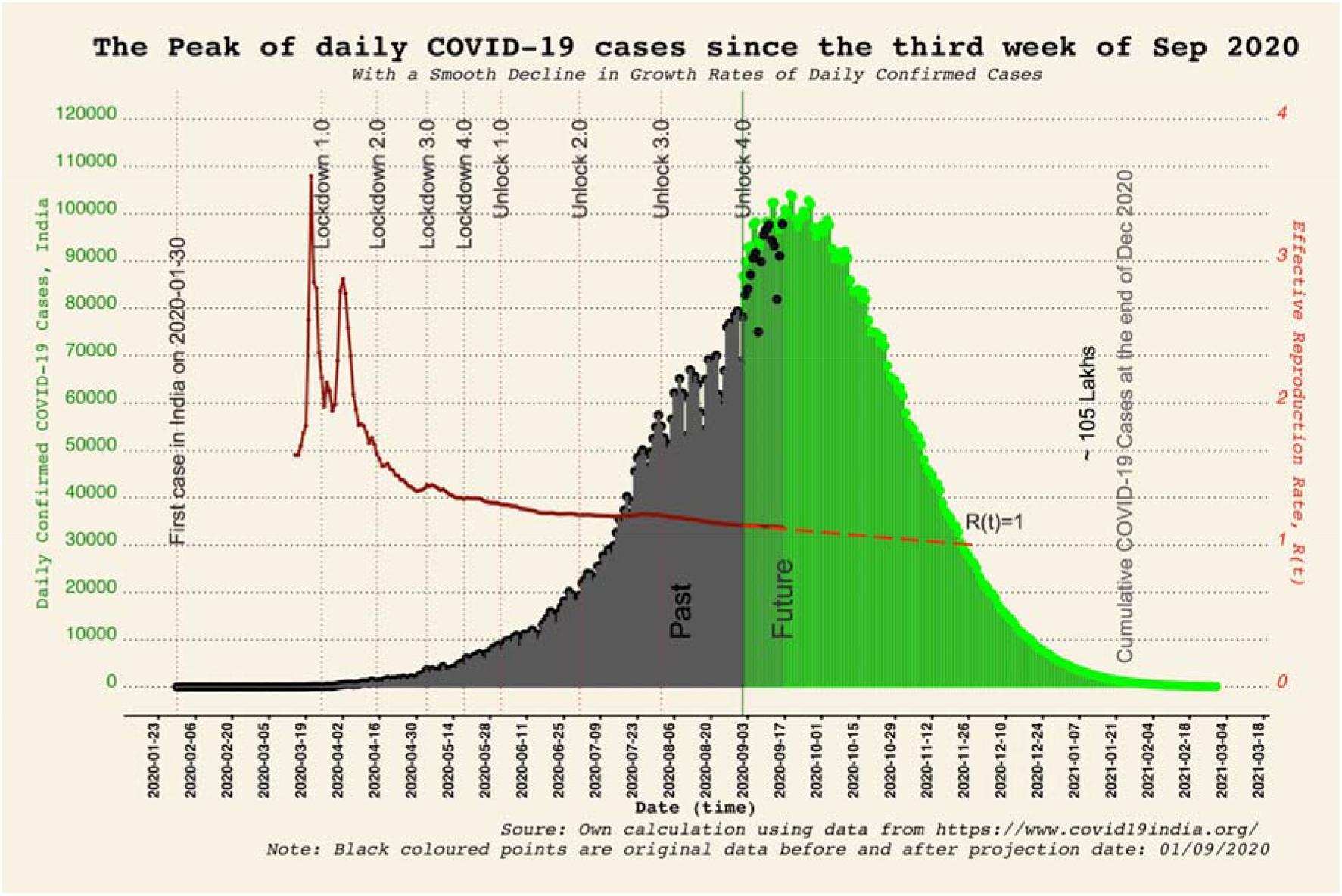
Projection of daily confirmed cases, India.

The *R*(*t*) values in India declined from the highest value of 3.6 persons in the third week of March to 1.4 persons at the end of April. During these two months of March and April, the trends in *R*(*t*) were non-steady. However, there was an increase in the *R*(*t*) values in the first week of May. The gradient of *R*(*t*) show an uptick. It was in resemblance to the ascent in growth rates. The growth rates have a significant effect on the *R*(*t*) as compared to serial interval distribution, and appropriately, the trends in *R*(*t*) is analogous to the trends in growth rates. The trends in *R*(*t*) were off the track of smooth decline from achieving low *R*(*t*) values in a shorter time as compared with the previous trends. The *R*(*t*) value was 1.3 at the end of May 2020, showing a marginal decline during these two-and-a-half months.

Nevertheless, the decline in the *R*(*t*) has been slow and timid during subsequent months from June to August 2020. It is apparent in the next two months of June and July when the trends in *R*(*t*) show almost near-stagnancy at the value of 1.21 persons. The warranted *R*(*t*) value of one is just a few points away but, given a length of time, slowly but surely, a bulk of confirmed cases were added in these two months. Nevertheless, in the next month of August, the *R*(*t*) values declined swiftly. During August, it declined from 1.21 persons in the first week to 1.14 persons at the end of the month. More importantly, a downward gradient of *R*(*t*) was evident after three months period since May 2020. This downward gradient would provide an expected *R*(*t*) values of one in near time.

### Projection, forecast and the peak of confirmed cases

For India, the projection of daily confirmed cases is the need of the hour, especially the forecast of the peak of daily confirmed cases. The projections for the growth rates were performed using the ARIMA model for obtaining the forecasts of daily confirmed cases. An ARIMA modelling procedure has four steps: assessment of model, estimation of parameters, diagnostic checking, and prediction. The time series plot of growth rates, autocorrelation function (ACF), and partial autocorrelation function (PACF) reveals seasonality and stationarity in growth rates of daily confirmed cases. ACF detects the degree of correlations between *y_t_* and y_t-k_ for different values of k, and PACF detects the degree of correlations between *y_t_* and *y_t-k_* conditional on *y_t-k 1_*,…, *y_t-1_*, i.e., partial correlations. Based on the ACF and PACF of the ARIMA model, the second difference was applied for making the time series stationary. The corrected AIC (AICc) value of the model ARIMA (2, 2, 1) was −347.3. Again, based on the seasonal part of ACF and PACF, the SARIMA (2, 2, 1) (1, 0, 1)[7] was applied that had the lowest AICc value of −357.3. The coefficients of this SARIMA model were significant at one per cent level (Table 1). The diagnosis of the developed SARIMA model confirms no trends in the residuals, no outliers, and nearly constant variance. Specifically, the ACF plot of residuals shows no significant autocorrelations. The plot of the residuals shows a standard normal variate. The p-values for the Ljung-Box statistic were above 0.05. In sum, the diagnosis of residuals confirms that the developed SARIMA (2, 2, 1) (1, 0, 1)[7] model is appropriate to the trends and seasonality in growth rates. Root mean square error (RMSE) and mean absolute percentage error (MAPE) of the SARIMA model were at 0.049 and 0.775, respectively, which is very small.

**Table 1:**
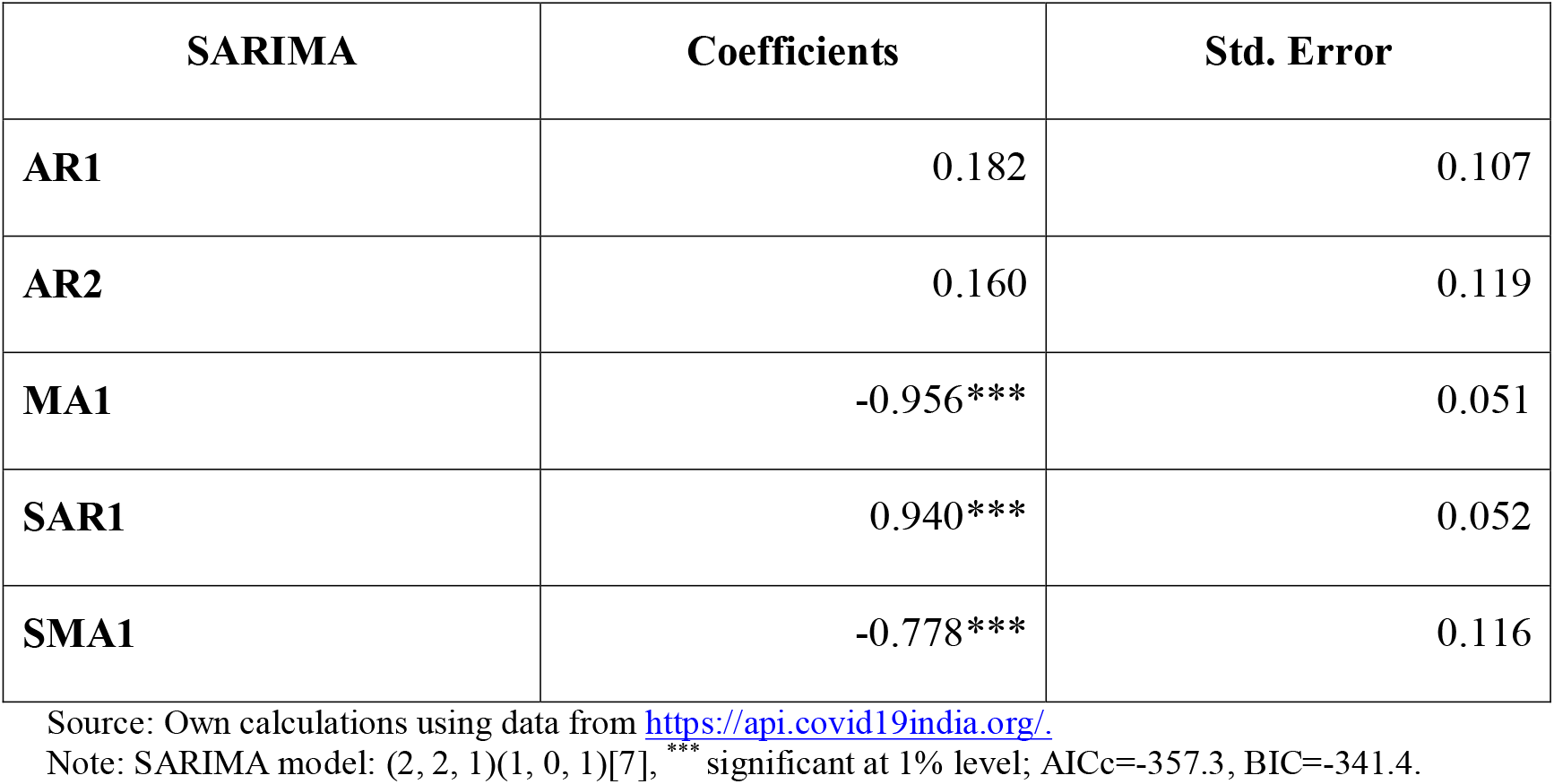
Parameters of SARIMA model.

| **SARIMA** | **Coefficients** | **Std. Error** |
| --- | --- | --- |
| **AR1** | 0.182 | 0.107 |
| **AR2** | 0.160 | 0.119 |
| **MA1** | -0.956*** | 0.051 |
| **SAR1** | 0.940*** | 0.052 |
| **SMA1** | -0.778*** | 0.116 |

Source: Own calculations using data from https://api.covid19india.org/.
Note: SARIMA model: (2, 2, 1)(1, 0, 1)[7], \*\*\* significant at 1% level; AICc=-357.3, BIC=-341.4.

Figure 3 shows the projected daily confirmed cases (green coloured) based on the SARIMA model from 1 September 2020 until 28 February 2021. The projected daily confirmed cases were nearly coinciding with the original data points between 1 September and 16 September. It ensures an independent cross-check compared to other sensitivity analysis based on past trends. Based on the projected growth rates, *R*(*t*) values were forecasted (long-dash red line).

The peak value of COVID-19 is the most warranted statistics among government authorities and academicians in India. The forecasted daily confirmed cases reveal normality around a peak. Given the pace of decline continues for the next month of September, the forecasts suggest the peak of confirmed cases since the third week of September. The forecast suggests that peak value would waver around 104,500 counts of daily confirmed cases in the third week of September. The projection reveals that the decline in the growth rates would not be steep. Therefore, a significant number of confirmed cases would be added yet witnessing the peak. The cumulative number of cases would reach a total of 105 lakhs by the end of December 2020. Sooner India observes a peak, lower the peak value and lesser the cumulative number of COVID-19 cases would be witnessed. However, the timing and magnitude of the peak to appear is contingent upon the swift decline of the growth rates in the next months, at the least in September 2020.

Figure 3 shows the *R*(*t*) values until it is equal to one (red coloured long-dash line). The *R*(*t*) values would be equal to one at a growth rate of value zero. However, the trends in growth rates suggest three months from September to reach a value of zero in the first week of December 2020. As per the forecast, it would take more time to go further low as compared to the decline of growth rates during the previous three months, i.e. between June and August. It is plausible because of the large base of confirmed cases after the peak shows up. The forecast suggests a significant lag between the peak value and the *R*(*t*) value of one.

For the Indian population, learning and practising social measures in the preceding seven months have ensured deceleration in the slope of the exponential model or a decline in the growth rates of daily confirmed cases. Nevertheless, it is important to recall the case of stagnant growth rates and *R*(*t*) values in June and July. India has shown a decline in the growth rates in August, and down the line, it is expected to maintain the pace of decline in September. At this point, a modest decline in growth rates would also be favourable to surpass the value of 2.0 and have lower growth rates between 2.0 and 1.0. Therefore, it is most expected to observe the peak in September 2020, which can happen in the third week of September 2020.

## Conclusion

The exponential model is the best fit over the daily confirmed COVID-19 cases during the last seven-and-a-half months in India. The applied exponential model reveals that initially, the growth rates of daily confirmed cases were high and non-steady positive values. However, the trends in growth rates show a linear decline during the five-and-a-half months period from the latter half of March to August 2020. Doubling time shows an increase in these five-and-a-half months period, concomitant with a steady decline in the growth rates. The *R*(*t*) values declined from the highest value of 3.6 persons in the mid-March 2020, 1.14 persons at the end of August 2020, to 1.12 persons on 16 September 2020. The trends in *R*(*t*) reveals that growth rates declined rapidly until April 2020 and after that, declined slowly until mid-September 2020. A slowdown in the *R*(*t*) is concomitant of a modest decline in the growth rates. The trends in *R*(*t*) show analogous to the trends in the growth rates of daily confirmed cases.

Based on the developed SARIMA model, the projection shows a smooth, slow decline in the growth rates of daily confirmed COVID-19 cases since September 2020. As per the projection, the growth rates would be less than 2.0 from the second week of September and would be one at the end of October and zero in the first week of December 2020. The forecast suggests the peak of the daily confirmed cases would appear since the third week of September. As per the forecast, the peak value would waver around 104,500 counts of daily confirmed cases since the latter half of September 2020. The *R*(*t*) value would be equal to one in the first week of December 2020. The total cumulative number of COVID-19 cases would be approximately 105 lakhs by the end of December 2020.

The study shows that the SARIMA model is suitable for projecting daily confirmed cases. This study based on moments of the distribution of the daily confirmed cases of COVID-19 disease unravels the uncertainty about the peak and curvature of COVID-19 disease. The study successfully explores the epidemiological stage of COVID-19 disease in India.

## Limitations of the study

The study is based on data of daily confirmed cases of COVID-19 disease during seven-and-a-half months between the dates 30 January 2020 and 31 August 2020. The data of future dates between 1 September and 16 September 2020 were used for validating the projections in the period studied. The API portal https://www.covid19india.org/ provides data on confirmed, active, deaths, and recovery cases; nevertheless, the data on daily confirmed cases is the most appropriate for projections. The exponential model applies to daily confirmed cases in India and its states. However, the projection analysis was performed at the national level only because smaller states do have limitations for projections because of small samples and differences in the length of period for COVID-19 disease.

The study is based on a few assumptions. Apart from statistical and time series assumptions, the behavioural aspects of population and socioeconomic and demographics characteristics of the population were not available for consideration in trend analysis and projections because of data constraints. The external factors such as capital and health expenditure were not possible to link with the time series. In the view of these data constraints, we do have trend and seasonality into the account for robust analysis and long-term projection.

## Data Availability

We retrieved data from an Application Programming Interface portal which is open access and publicly available.

https://www.covid19india.org/

## Acknowledgements

Authors would like to thank Prof. K. S. James, the Director and Senior Professor, International Institute for Population Sciences (IIPS), Mumbai, for his recommendations and suggestions to carry out this study, initiating policy briefs on COVID–19, and for providing constructive comments which helped greatly in improving the quality of this work. We are also thankful to the members of the group “IIPS Team of Researchers on Estimation and Projection of COVID 19 Cases” for their suggestions.

